# Fine adaptive control of precision grip without maximum pinch strength changes after median nerve mobilization

**DOI:** 10.1101/2020.04.01.20049635

**Authors:** Frédéric Dierick, Jean-Michel Brismée, Olivier White, Anne-France Bouché, Céline Périchon, Nastasia Filoni, Vincent Barvaux, Fabien Buisseret

## Abstract

Fine dexterity critically depends on information conveyed by the median nerve. While the effects of its compression and vibration are well characterized, little is known about longitudinal tension and excursion. Using a force-sensitive manipulandum, a numeric dynamometer and Semmes-Weinstein monofilaments, we examined the adaptations of precision grip control, maximum pinch strength and fingertips pressure sensation threshold before and immediately after the application of longitudinal tension and excursion mobilizations applied on the median nerve. Grip (*G*_*F*_) and load (*L*_*F*_) forces applied by the thumb, index and major fingers were collected in 40 healthy young participants during three different grip precision tasks along the direction of gravity. For grip-lift-drop task, maximum *G*_*F*_ and *L*_*F*_ and their first time derivatives were computed. For up-down oscillations, means of *G*_*F*_ and *L*_*F*_ and their variability were computed. For oscillations with up and down collisions, peaks of *G*_*F*_ and *L*_*F*_, time delay between *G*_*F*_ peak and contact, and values of *G*_*F*_ and *L*_*F*_ at contact were collected. Our findings show that median nerve mobilizations induce significant fine adaptations of precision grip control in the three different tasks but mainly during grip-lift-drop and oscillations with collisions. Fingertips pressure sensation thresholds at index and thumb were significantly reduced after the mobilizations. No significant changes were observed for maximum pinch strength. We conclude that precision grip adaptations observed after median mobilizations could be partly explained by changes in cutaneous median-nerve mechanoreceptive afferents from the thumb and index fingertips.

## Introduction

The median nerve is one of the major mixed peripheral nerves of the upper limb and a key player in hand function, in particular during the control of fine object manipulation, both at the sensory and motor levels^1–5^. In fact, during dexterous manipulation, there is a close interplay between sensory mechanisms related to explorative functions of the fingers and motor mechanism controlling the muscles of the hand^6^.

For the past 30 years, lessons about the role of the median nerve in precision grip control have been documented in studies in healthy subjects using the microneurography technique^1^, or anesthetic blocks of the nerve at the level of the wrist^5, 7^ or hand^6, 8–13^. All showed that the median nerve plays an important role in scaling the grip force and coupling of grip force with load forces when lifting, holding or oscillating an object^7, 8, 10–12, 14^, by regulating the timing and amplitude of grip force development or the adaptation to perturbation^6, 8, 9, 11, 14^. Similar conclusions were made when generating torques of force production patterns^5, 11^.

A functional grip force control is critical in everyday life. Therefore, the role of the median nerve in precision grip control has been also specified in clinical studies including patients suffering from chronic entrapment of the median nerve at the wrist, also known as carpal tunnel syndrome^2, 15–18^. Additionally, active median nerve self-mobilizations or passive mobilizations are frequently prescribed or executed by orthopaedic manual physical therapists with the aim of restoring the physiological function of the nerve. Nerve mobilization reduces intra-neural pressure, leading to increased capillary blood flow and oxygen supply to the nerves. Hence, this mechanism improves axoplasmic flow and, consequently, nerve conduction^19^.

Even if very few studies explored the relationships between median nerve conduction velocity and grip, it should be noted that median nerve conduction velocity contributes to maximum grip strength in healthy subjects^20^ and correlates to accuracy in precision grip force control in recurrent carpal tunnel syndrome patients^21^. Previous studies observed that median nerve self-mobilization or applied by an orthopaedic manual physical therapist increase nerve conduction velocity^22^, specifically in the elbow-to-wrist section and in closed kinetic chain condition^23^. Even if median nerve neural mobilization was not effective to produce increase in maximum grip strength in healthy subjects^24^, the assessment of 3-jaw chuck pinch strength, that allows better targeting of the median nerve, has never been explored after its mobilization. In a rather logical way, the median nerve is also involved in maximum pinch strength generation^17, 25, 26^. Furthermore, in patients with carpal tunnel syndrome, maximum pinch strength was significantly increased after median nerve mobilization exercises^27, 28^. These results could be explained by induced fluid dispersion in the median nerve at the carpal tunnel following its mobilization^29^ and a decrease in fingertips pressure sensation threshold after median nerve mobilization exercises^28^.

Among mechanical stimuli applied to the median nerve, the impact of compression and vibration on precision grip have been mainly explored, but the effects of longitudinal tension or excursion remains unclear even today. The main objective of this study was to examine the physiological grip responses in three different precision tasks, before and immediately after the application of median nerve mobilizations. We adopted these tasks because they contain control components that are highly relevant in myriad of functional activities and should therefore reflect a healthy control. Concomitantly, maximum 3-jaw chuck pinch strength and fingertips pressure sensation thresholds at thumb, index, and major fingers were assessed to attempt explaining the potential changes in precision grip responses.

## Methods

### Participants

Forty-nine healthy undergraduate and graduate students were recruited from our Physical Therapy Department at Haute Ecole Louvain en Hainaut and 40 (24 males/16 females; age: 26±2 years) participated in this trial. Participants did not receive financial compensation for their participation. As inclusion criteria, participants were aged between 18 and 30 years, and not currently experiencing neither neck nor dominant upper extremity symptoms. Participants were excluded (n=9) if they had a Disabilities of the Arm, Shoulder and Hand (DASH)^30^ score larger than 1. Thirty-four participants were right-handed and six were left-handed, according to the Edinburgh inventory^31^.

The participants received randomly an upper limb neurodynamic test (ULNT1) mobilization intended to provide anatomical stress across the median nerve based on either a tensioning maneuver (*n* = 20) or a sliding maneuver (*n* = 20). All participants were naïve to the experimental objectives. The study protocol and the informed consent documents were approved by the Academic Ethical Committee “Brussels Alliance for Research and Higher Education”. All research was performed in accordance with relevant guidelines/regulations, and informed consent was obtained from all participants.

### ULNT1 maneuvers

An ULNT1 tensioning maneuver is obtained by moving one or several joints in such a manner that the nerve bed is elongated^32^. The tensioning maneuver included the following movements: shoulder depression, abduction and external rotation to 90 degrees, full elbow extension, and forearm supination, and contralateral cervical spine side bending. An ULNT1 sliding maneuver consists of an alternation of combined movements of at least two joints in which one movement lengthens the nerve bed thus increasing tension on the nerve while the other movement simultaneously decreases the length of the nerve bed which unloads the nerve^32^. The sliding maneuver included the following movements: shoulder depression, abduction and external rotation to 90 degrees, full elbow extension, and forearm supination, and ipsilateral cervical spine side bending. For ULNT1 tensioning and sliding maneuvers, a licensed orthopaedic manual physical therapist (F.D.) performed the upper quadrant mobilization while another one (C.P. or N.F.) the passive mobilization of the cervical spine. To standardize the dosage, twenty repetitions were performed from full wrist and finger flexion to full wrist and finger extension and back.

### Pinch strength

A 3-jaw chuck pinch, also known as palmar pinch, is a pinch technique that involves placing the thumb on one side of the pinchmeter and the index and major fingers on the other side^33^. The maximum voluntary pinch strength of the dominant hand was determined with a numeric pinchmeter (P100, Biometrics Ltd., Gwent, UK) while the participant sat with shoulder adducted to his/her side and neutrally rotated, elbow flexed to 90°, forearm in a neutral position, and the wrist held between 0-15° of ulnar deviation. The gauge displayed values in kg with the range of values being 0 to 22 kg. Maximum force was determined three times by the same examiner (C.P.), before the first precision grip control assessment and after the second one. The mean of 3 measurements was calculated and used for data analysis^33^. Before measurements, the examiner demonstrated the correct placement of the fingers on the dynamometer, and standardized verbal instructions^33^ were given to each participant.

### Fingertips pressure sensation threshold

Semmes-Weinstein monofilament (SWM) testing was conducted by the same examiner (N.F.), before the first precision grip control assessment and after the second one. Participants closed their eyes and were instructed to verbally indicate when they felt the monofilament. Pressure was applied to the fingertips of thumb, index and major fingers to just bend the monofilament and each individual stimulus was applied up to 3 times. The smallest monofilament at which the participant indicated sensation was recorded as the threshold.

### Precision grip control assessments

#### Apparatus and data processing

The apparatus (GLM-Box, Arsalis, Belgium) used for these experiments was a grip-lift manipulandum (GLM) composed of different mechanical parts made of an aluminum 7075 alloy, assembled with screws (Fig. 1). The mass of the GLM was 0.262 kg and its dimensions (height × width × depth) were 91 × 37.5 × 48 mm. The GLM is made of two half hollow shells supporting a grip circular surface (diameter: 40 mm) covered with brass. Each gripping surface is equipped a three-dimensional force-torque sensor (Mini40 F/T transducer; ATI Industrial Automation, Apex, NC, USA). An additional tridimensional accelerometer (Analog Devices, ref. ADXL330) and a printed circuit board are embedded inside the shells. The sensors measured the forces applied to the centres of the grasp surfaces and recorded the components (*F*_*x*_, *F*_*y*_, *F*_*z*_) in the cartesian frame defined in Fig. 1. Sensing ranges for *F*_*x*_, *F*_*y*_, and *F*_*z*_ were 40, 40, and 120 N, with 0.01, 0.01, and 0.02 N nominal resolution, respectively. Analog signals were amplified, filtered with a Bessel 150-Hz cut-off low-pass fourth-order filter, and sampled at 1 kHz.

**Figure 1.**
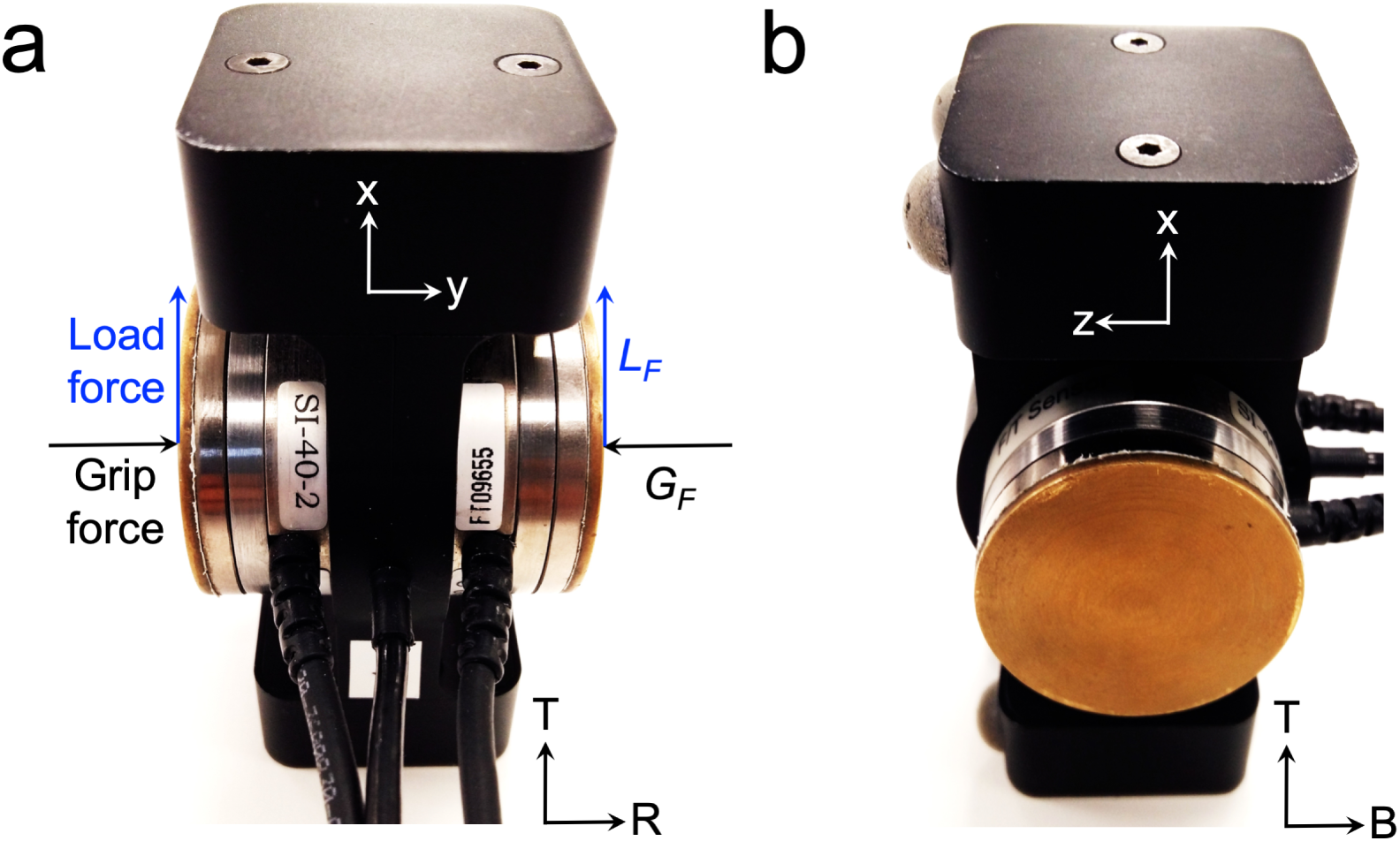
(a) Rear view of the GLM. Schematic representation of total grip (*G*_*F*_, in black) and load vector forces (*L*_*F*_, in blue) applied on right (thumb) and left (index and major) sensors. T: top; R: right side (b) Side view (left) of the GLM. T: top; B: back. The cartesian frame (x,y,z) used to compute *G*_*F*_ and *L*_*F*_ is shown on the two different views of the GLM.

Data were analyzed offline. The total grip force magnitude (*G*_*F*_) was calculated as the average magnitude of the normal forces *F*_*y*_ applied by the thumb and the index-major fingers on the right (*F*_*y,r*_) and left (*F*_*y,l*_) sensors, respectively:

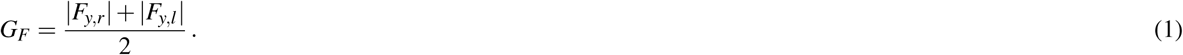

In equation (1), *F*_*y,r*_ and *F*_*y,l*_ are the normal force components on the right and left sensors, respectively.

The total load force (*L*_*F*_) magnitude was defined as the sum of the right and left tangential forces and computed as follows:

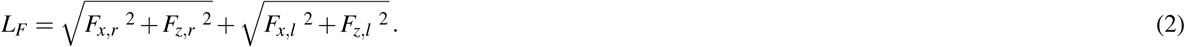

In equation (2), *F*_*x*_ and *F*_*z*_ are the horizontal and vertical force components of the tangential force applied on the right (*r* subscript) and left (*l* subscript) sensors.

#### Procedure

Five minutes before the experiments, participants washed their hands with soap and water and were allowed to familiarize themselves with the physical characteristics of the apparatus by handling it. Each participant sat in a height-adjustable chair without armrests in front of a desk supporting the apparatus. Her/his shoulders were slightly abducted, hands resting on the horizontal surface of the desk, hip flexed around 90 degrees, lower back supported by backrest, and feet flat on the ground. At a signal from the experimenter, they were instructed to keep their non-dominant hand at rest, and to pick up the manipulandum using a precision grip between the thumb and the index-major fingers of their dominant hand. The experiment comprised three different grip precision tasks along the direction of gravity that are described hereafter. In all tasks, participants were instructed to move the apparatus vertically and to keep its orientation constant during the movement. The time elapsed between the end of median nerve mobilizations and the end of the last precision grip task was about 3–4 minutes. All participants were familiarized with the three grip precision tasks described hereafter.

#### Grip precision tasks

In the first task, participants were instructed to perform grip-lift-drop movements6, during 30 seconds. Participants were instructed to lift the apparatus up to about 30 cm above the desk and to keep it about this position for around 2-3 s, and then replace it gently on the desk. Only the grip-lift phase was analyzed and the lifting movement took place mainly as a radial deviation of the wrist, a flexion of the elbow and the shoulder. Outcome measures for the grip-lift phase included: maximum *G*_*F*_ (*G*_*F*_ max) and *L*_*F*_ (*L*_*F*_ max) and the maximum of their first time-derivatives of the force signals (d*G*_*F*_ max and d*L*_*F*_ max) that were computed by ±5-point numerical differentiation. These parameters are illustrated in Fig. 2a and b.

**Figure 2.**
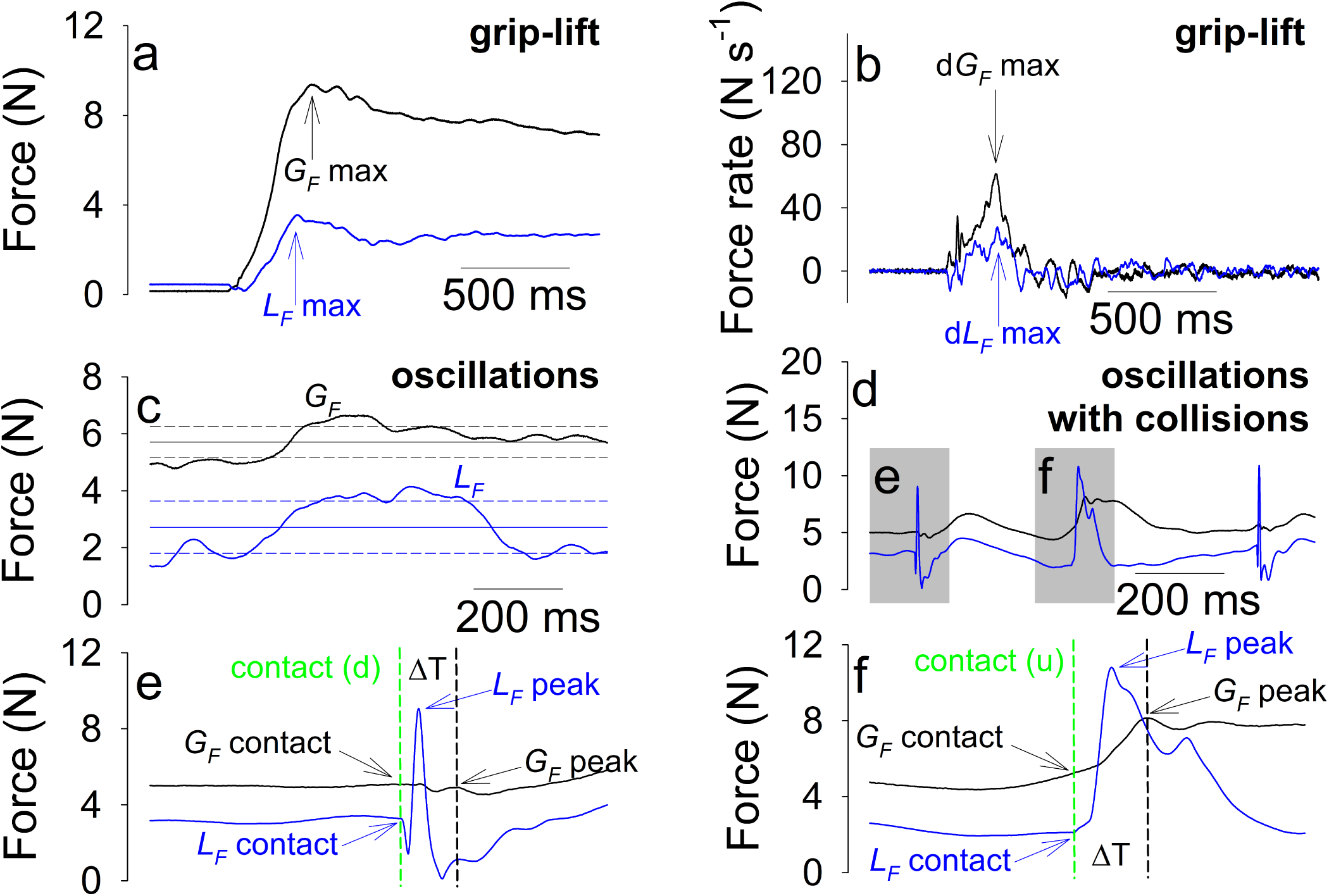
(a) Typical time traces of *G*_*F*_ (in black) and *L*_*F*_ (in blue) variables and parameters (*G*_*F*_ max and *L*_*F*_ max) collected during the grip-lift phase of the grip-lift-drop movement; (b) Typical time traces of d*G*_*F*_ (in black) and d*L*_*F*_ (in blue) variables and parameters (d*G*_*F*_ max and d*L*_*F*_ max) collected during the grip-lift phase of the grip-lift-drop movement; (c) Typical time traces of *G*_*F*_ (in black) and *L*_*F*_ (in blue) variables and parameters (*G*_*F*_ mean [black horizontal line], *G*_*F*_ SD [black horizontal short dash lines], *L*_*F*_ mean [blue horizontal line], and *L*_*F*_ SD [blue horizontal short dash lines]) collected during the vertical oscillations; (d) typical traces of *G*_*F*_ and *L*_*F*_ variables collected during vertical oscillations with up (inset e) and down (inset f) collisions; (e) Parameters (*G*_*F*_ contact, *G*_*F*_ peak, *L*_*F*_ contact, *L*_*F*_ peak, and ΔT) collected during the upward collisions (u); Parameters (*G*_*F*_ contact, *G*_*F*_ peak, *L*_*F*_ contact, *L*_*F*_ peak, and ΔT) collected during the downward collisions (d). Traces were collected in a 26-year-old right-handed female participant.

In the second task, the participants were instructed to perform vertical rhythmic arm movements above or below the hand’s neutral position between two targets positioned 30 cm apart, during 30 seconds. The oscillations were timed by a metronome at a frequency of 1 Hz per up-down cycle. The oscillatory movements took place mainly as a flexion-extension of the elbow and the shoulder. Outcome measures for oscillations included: mean of *G*_*F*_ and *L*_*F*_ (*G*_*F*_ mean and *L*_*F*_ mean) and their variability (standard deviation, SD, *G*_*F*_ SD and *L*_*F*_ SD). These parameters are presented in Fig. 2c.

In the third task, the participants were instructed to perform 30 cm vertical rhythmic arm movements and tap a target situated above or below the hand’s neutral position with the GLM, during 30 seconds. In this task, an interaction with the environment had to be planned as well, similar to laying down a mug of coffee on a table or hanging an object on a hook. The oscillations with up and down collisions were also timed by a metronome at the same frequency as in the previous task. The oscillation movements with collisions took place mainly as a flexion-extension of the elbow and the shoulder. Outcome measures for oscillations movements with collisions included: peaks of *G*_*F*_ and *L*_*F*_ (*G*_*F*_ peak and *L*_*F*_ peak) and values of *G*_*F*_ and *L*_*F*_, 1 ms before target contact (*G*_*F*_ contact and *L*_*F*_ contact)34. Time delay between *G*_*F*_ peak and contact was computed as ΔT. These variables were separately recorded for the up and down phases and contacts. These parameters are presented in Fig. 2d to f.

These three tasks all required fine control of the hand while processing the dynamics of the object in the environment by the sensorimotor system. The dynamics of the object is predominant during grip-lift-drop, while the dynamics of the upper limb is predominant during oscillations without collisions. During oscillations with collisions, the dynamics of both object (during collisions) and upper limb (during upward and downward displacement of the object) must be taken into account. In addition, grip-lift-drop is a discrete task, whereas the oscillations, with or without collisions, are rhythmic. In order to be able to perform these tasks without dropping the object, the sensorimotor system must have formed and shared internal models of the dynamics that capture the mechanical behavior of the object while interacting with the hand.

## Statistical analysis

Data were assessed for normality (Shapiro-Wilk) and equal variance (Brown-Forsythe) tests. One-way (time) repeated measures (RM) Analysis of Variance (ANOVA) tests were used for variables related to the pinch strength and the grip precision tasks assessments, before and after the median nerve mobilizations (time factor). Two-way (time×location) RM ANOVA tests were used for variables related to the fingertips pressure sensation threshold assessments at the thumb, index, and major (location ±factor), and before and after the median nerve mobilizations (time factor). These tests were followed by Holm-Sidak post hoc for multiple-comparisons testing when significant results were observed. The significance level was set at *p* = 0.05 for all analyses. Data are presented as mean±SD. All statistical procedures were performed with SigmaPlot software (version 13.0, Systat Software, San Jose, CA).

## Results

Typical time traces of *G*_*F*_ and *L*_*F*_ recorded before and after the median nerve mobilizations during the grip-lift-drop task are shown in Fig. 3a. *L*_*F*_ value during stationary holding of the object between each movement was equal to the object’s weight (2.57 N, Fig. 3a). Significant increases of *L*_*F*_ max, d*G*_*F*_ max and d*L*_*F*_ max were observed after the mobilizations (Table 1).

**Table 1.** One-way RM ANOVA results before and after mobilizations for for all parameters collected during the pinch strength and the grip precision tasks.

| Variable | Before (Mean±SD) | After (Mean±SD) | F value | p value |
| --- | --- | --- | --- | --- |
| **Pinch strength** |  |  |  |  |
| Maximum force (kg) | 8.1±2.0 | 8.4±2.2 | 2.12 | 0.153 |
| **Grip-lift-drop** |  |  |  |  |
| $G_F$ max (N) | 15.2±13.4 | 15.0±11.1 | 0.03 | 0.86 |
| $L_F$ max (N) | 3.2±0.4 | 3.4±0.4 | 6.66 | **0.014** |
| $dG_F$ max (N s<sup>-1</sup>) | 89.0±66.6 | 106.2±59.6 | 7.54 | **0.009** |
| $dL_F$ max (N s<sup>-1</sup>) | 43.6±17.0 | 56.0±17.9 | 19.56 | **<0.001** |
| **Oscillations** |  |  |  |  |
| $G_F$ mean (N) | 8.1±4.0 | 8.1±4.7 | 0.001 | 0.974 |
| $L_F$ mean (N) | 2.3±0.2 | 2.4±0.3 | 2.59 | 0.116 |
| $G_F$ SD (N) | 1.8±1.5 | 1.8±1.6 | 0.004 | 0.951 |
| $L_F$ SD (N) | 0.9±0.3 | 1.0±0.2 | 9.34 | **0.004** |
| **Oscillations with collisions (up)** |  |  |  |  |
| $G_F$ peak (N) | 13.3±7.1 | 12.5±7.3 | 1.39 | 0.245 |
| $L_F$ peak (N) | 17.4±8.3 | 15.1±7.5 | 15.35 | **<0.001** |
| $G_F$ contact (N) | 12.4±6.7 | 11.3±6.8 | 4.88 | **0.033** |
| $L_F$ contact (N) | 2.9±0.4 | 3.00±0.4 | 6.14 | **0.018** |
| $\Delta T$ (ms) | 74.9±39.8 | 74.6±32.9 | 0.003 | 0.956 |
| **Oscillations with collisions (down)** |  |  |  |  |
| $G_F$ peak (N) | 13.5±7.4 | 12.3±7.7 | 5.05 | **0.030** |
| $L_F$ peak (N) | 14.5±6.0 | 13.6±5.5 | 6.11 | **0.018** |
| $G_F$ contact (N) | 11.7±6.7 | 10.5±6.8 | 3.02 | 0.090 |
| $L_F$ contact (N) | 2.3±0.8 | 2.4±0.9 | 0.128 | 0.722 |
| $\Delta T$ (ms) | 45.4±30.4 | 46.8±30.3 | 0.212 | 0.648 |

**Figure 3.**
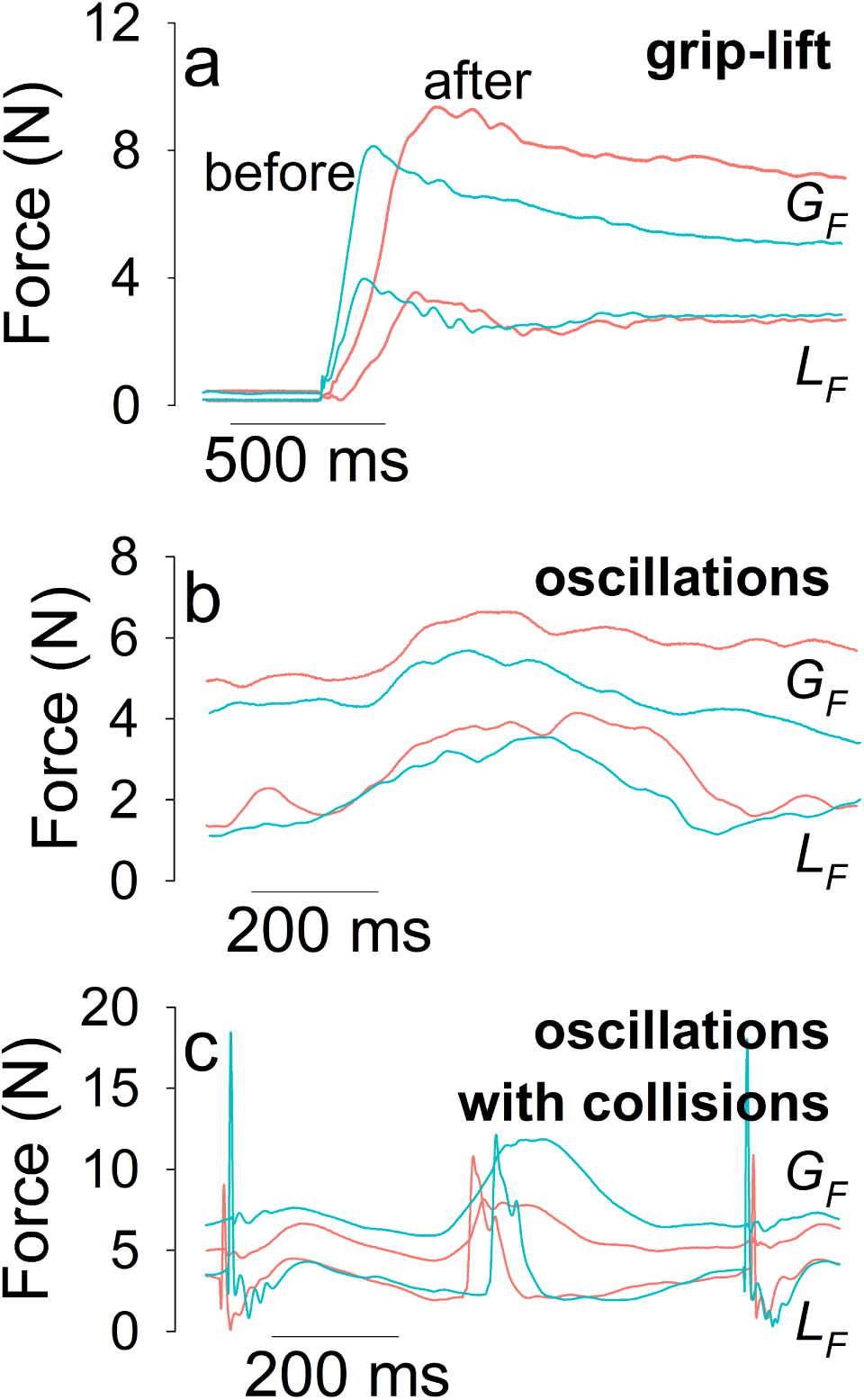
(a) Typical time traces of *G*_*F*_ and *L*_*F*_ variables during the grip-lift phase of the grip-lift-drop movement, before (turquoise) and after (pink) the mobilizations; (b) typical time traces of *G*_*F*_ and *L*_*F*_ variables during the vertical oscillations, before (turquoise) and after (pink) the mobilizations; (c) typical time traces of *G*_*F*_ and *L*_*F*_ variables during the vertical oscillations with collisions, before (turquoise) and after (pink) the mobilizations. Traces were collected in the same participant as in Fig. 2.

Typical time traces of *G*_*F*_ and *L*_*F*_ variables recorded during the vertical oscillation tasks, without and with collisions, are presented in Fig. 3b and c, respectively. A significant increase of *L*_*F*_ SD was observed after the mobilizations during the oscillations without collisions (Table 1), indicating a greater variability of the amount of *L*_*F*_ modulation. A significant decrease of *L*_*F*_ peak and *G*_*F*_ contact and increase of *L*_*F*_ contact were observed after the mobilizations during oscillations with collisions in upward direction (Table 1).

A significant decrease of *G*_*F*_ peak and *L*_*F*_ peak were observed after the mobilizations during oscillations with collisions in downward direction (Table 1). Split violins plots also indicating mean results obtained for each parameters are presented in Fig. 4 for grip-lift and oscillations without collisions and in Fig. 5 for oscillations with collisions. SWM testing results are shown in Fig. 6. Multiple comparison testing showed that fingertips pressure sensation threshold at index (*t* = 3.314, *p* = 0.001) and thumb (*t* = 2.716, *p* = 0.008) were significantly reduced after the mobilizations, indicated by lower means for number of SWM (Fig. 6). Results for maximum force recorded during the pinch strength assessment are shown in Table 1. No significant difference was observed after the mobilizations.

**Figure 4.**
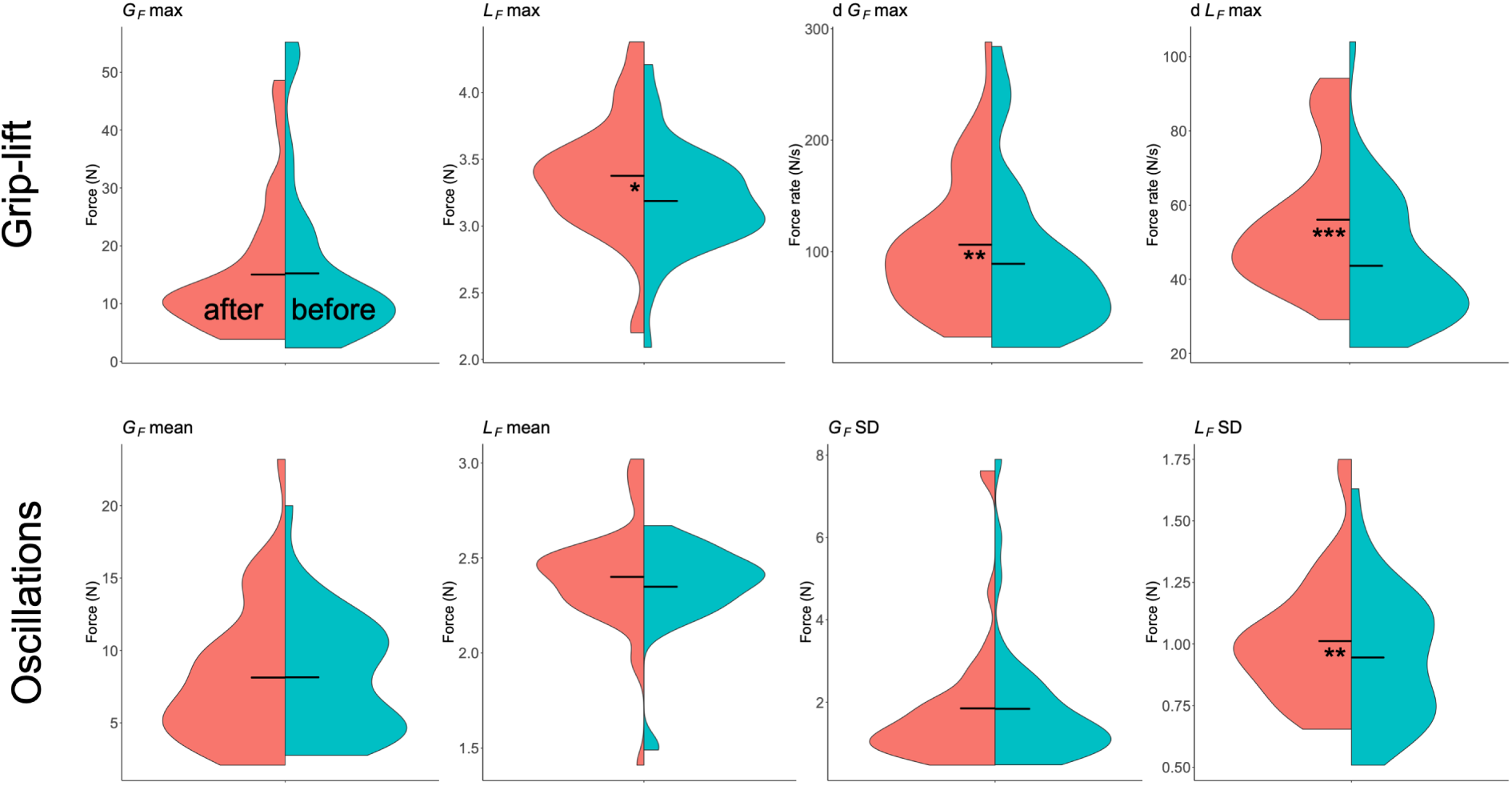
Split violin plots for all parameters computed before (turquoise) and after (pink) median nerve mobilizations during grip-lift (upper row) and oscillations without collisions (lower row). Horizontal bars represent mean values. Significant differences are specified with stars.

**Figure 5.**
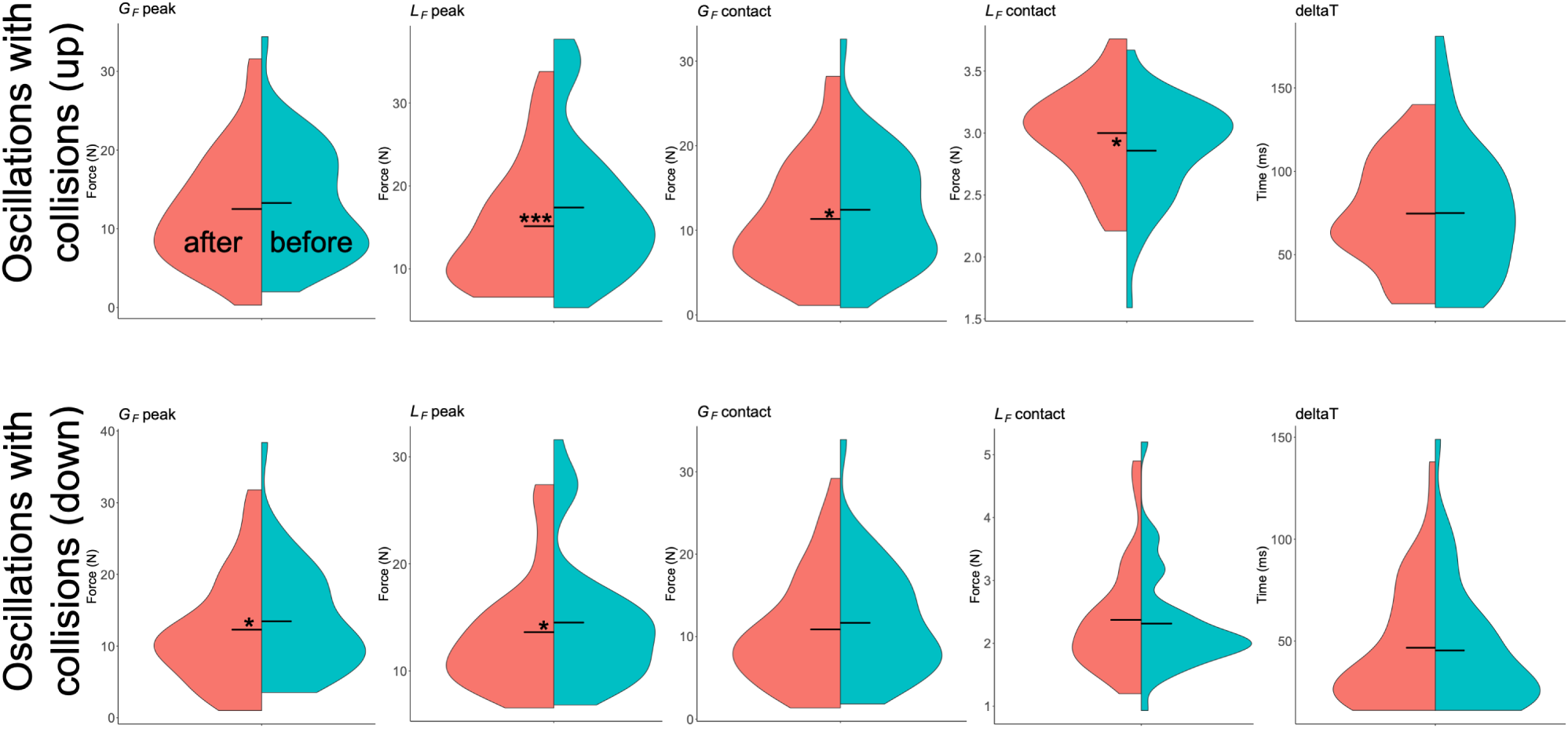
Split violin plots for all parameters computed before (turquoise) and after (pink) median nerve mobilizations during oscillations with upward collisions (upper row) and downward collisions (lower row). Horizontal bars represent mean values. Significant differences are specified with stars.

**Figure 6.**
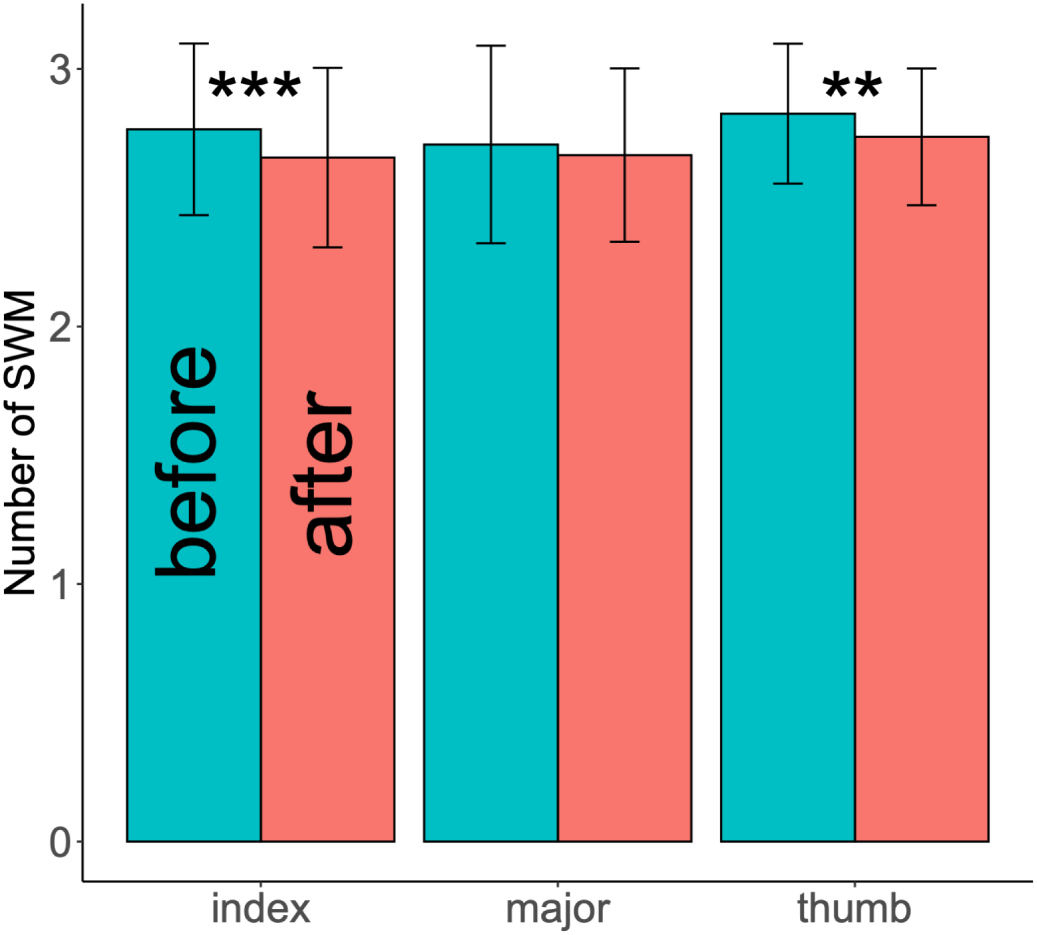
Bar chart with mean and SD results for SWM testing results, before (turquoise) and after (pink) mobilizations at the fingertips of thumb, index and major. Significant differences were observed for thumb and index and are specified with stars.

## Discussion

This study intended to capture the effects of median nerve mobilizations on precision grip control during grip-lift-drop, oscillations and a mixed task involving transport of an object with collisions. All participants succeeded in completing the tasks without dropping the object. Since orthopaedic manual physical therapists use median nerve mobilizations to treat patients with neck and upper limb pain conditions, understanding the effects of this intervention on precision grip control is of major importance. Before investigating patients with pain, it is relevant to assess the physiological responses in a sample of healthy subjects. Identification of physiological grip responses to median nerve mobilizations is very useful for identifying side effects of the latter on the safety of manipulating objects and will facilitate the identification of symptomatic subjects’ abnormal responses. In addition to parameters computed from grip and load forces applied by the thumb, index and major fingers, maximum pinch strength and fingertips pressure sensation thresholds were assessed. Our findings show that median nerve mobilizations induces significant adaptations of precision grip control in the three different tasks but mainly during grip-lift-drop and oscillations with collisions. At the same time, pressure sensation thresholds at index and thumb were significantly reduced, without changes in maximum pinch strength.

The main strength of this study is that it has explored three motor tasks involving the sensorimotor system in different ways, either predominantly at the level of the dynamics of the object, the upper limb or both. From a motor control point of view, these tasks are supported both by feedforward and feedback mechanisms used by the central nervous system. Our hypothesis is that the mobilizations would affect the feedback mechanisms from the median nerve, which could be compensated by internal models. These different tasks revealed fine physiological modifications of the precision grip, mainly at the level of force applied tangentially to the fingertips (*L*_*F*_).

In the grip-lift-drop task, the pattern of *G*_*F*_ adjustments to object-induced *L*_*F*_ fluctuations is consistent with those observed in previous studies^1, 6^. *G*_*F*_ max was not changed after the mobilizations. Since this parameter reflects the capacity of the motor system to process incoming sensory information signaling of object weight from lift-off to update motor output accordingly^35–37^, we conclude that this sensory process related to the object’s weight was not altered after the mobilizations. However, d*G*_*F*_ max was increased, which means that the grip rate performed by the participants is faster after the mobilizations. The d*G*_*F*_ max before lift-off is the most sensitive measure to indicate successful preplanning of manual interaction with familiar objects^35–37^ and provides an index that quantifies how fast muscle fibers can be recruited. Since during our experimental procedure all participants were allowed to familiarize with the object before the measurements, we can conclude that the mobilizations altered this preplanning phase. Muscle responses were also faster.

An increase in *L*_*F*_ max and d*L*_*F*_ max was also observed during the grip-lift-drop task, which means that the lifting movements performed by the participants were faster after the mobilizations. Faster movements to lift objects were previously observed in elderly people and interpreted as a strategy of relying predominantly on feedforward control in order to compensate for less functional afferent feedback^38^. Regardless of the effect of aging, we believe that the adoption of a similar strategy in our young participants is possible. From a kinematic point of view, lifting of the object is mainly related to radial deviation of the wrist and flexion of the elbow and shoulder. It is therefore interesting to ask to what extent the median nerve can be involved in controlling wrist, elbow, and shoulder movements. Wrist radial deviation is mainly controlled by *flexor carpi radialis* and *extensor carpi radialis longus* muscles. The latter is innervated by the radial nerve while the former is innervated by the median nerve. Flexion of the elbow and shoulder are controlled by the musculocutaneous nerve that innervates *coracobrachialis, biceps brachii*, and *brachialis* muscles. However, anastomoses between the musculocutaneous and median nerves in the arm were seen in more than 507 of adult and fetuses cadavers^39^, suggesting that the median nerve can almost partly contribute to the flexion movements of the shoulder and elbow.

During up and down oscillations, the pattern of *G*_*F*_ adjustments to the movement-induced *L*_*F*_ fluctuations is consistent with those observed in previous studies^40–43^. The only significant change after the mobilizations was an increase of *L*_*F*_ SD. It is reasonable to speculate that the increased *L*_*F*_ variability indicates excessive noise in the sensorimotor system of the participants to control instabilities linked to upper limb kinematics. This excessive noise observed at the fingertips could reflect the decrease in kinematic performance. However, variability in a physiological process is thought to be necessary to adapt to unpredictable environmental changes^44^ and it is now necessary to study this variability with other indicators than standard deviation like fractal tools for example. Fractal analysis would allow to study not only the variability’s magnitude (SD) but also its dynamical structure; we leave such a study for future works.

During oscillations with collisions, regardless of the direction, *L*_*F*_ peak was decreased after the mobilizations. *L*_*F*_ peaks appeared just after contact. The decrease of *L*_*F*_ peak means that the participants were voluntarily hitting the targets less hard, probably to prevent the object from slipping. Consequently to *L*_*F*_ peak decrease, *G*_*F*_ peak also decreased in the two directions but it was only significant in the downward direction. Moreover, in upward direction, *G*_*F*_ contact was significantly reduced after the mobilizations while *L*_*F*_ contact was increased. Note also that *G*_*F*_ contact was almost significantly changed in downward direction. A reduced *G*_*F*_ contact value is advantageous to rapidly absorb the vibrations at the beginning of the collisions before increasing *G*_*F*_ to a maximum (*G*_*F*_ peak)^34^ to restore stability. Since *L*_*F*_ contact value is related to the acceleration of the object during the transport phase, its increase after the mobilizations would be related to kinematic changes in the upper limb, as previously discussed for the grip-lift-drop task.

Pressure sensation thresholds at index and thumb fingertips were significantly reduced after the mobilizations, indicating changes in cutaneous median-nerve mechanoreceptive afferents. At this stage of knowledge, our hypothesis is that the increase in median nerve conduction velocity that has been observed after mobilizations in previous studies^22, 23^ is implicated in such change.

Various limitations must be acknowledged. First, the changes could be shown by carrying out the measurements immediately after the mobilizations and probably do not last beyond the time required for the completion of the three tasks. Further research should investigate the length of time these changes would last. Second, we do not include a control group without median nerve mobilizations. Third, tensioning and sliding mobilization results were lumped together since no specific “mobilization type” effect was observed using a two-way RM ANOVA. Fourth, we do not know yet the clinical meaningfulness of pressure and precision grip changes. Nevertheless, physiological changes in precision grip control as a result of median nerve mobilization have been demonstrated for the first time.

## Data Availability

All data are available.

## Acknowledgements

With the financial support of the European Regional Development Fund (Interreg FWVl NOMADe).

## Author contributions statement

F.D. conceived and conducted the experiments and analyzed the results. C.P. conducted the experiments and analyzed the results. N.F. conducted the experiments and analyzed the results. O.W. conceived the experiments and analyzed the results.

All authors reviewed the manuscript.

## Additional information

The authors declare no competing financial/ or non-financial interests.

